# Disease-modifying therapies for T1 hypointense lesions (Black Holes) volume in patients with multiple sclerosis: Protocol for a systematic review and meta-analysis

**DOI:** 10.1101/2021.06.25.21259388

**Authors:** Mohammad Reza Fattahi, Mohammad Ali Sahraian, Amir Reza Azimi, Maryam Sadeghi, Mehrnush Torbati, Amir Valizadeh

## Abstract

**Rationale:** A major part of diagnosis and follow-up in patients with multiple sclerosis is based on MRI evaluations. As T1-hypointense lesions represent neural destruction or axonal loss and anticipate irreversible disability in the patients, evaluation of the effects of Disease-modifying therapies (DMTs) on the volume of black holes seems necessary.

**Objectives:** To evaluate the effects of FDA approved disease-modifying therapies (DMTs) on T1 hypointense lesions (Black Holes) volume in patients with multiple sclerosis (MS)

**Data sources:** We will search MEDLINE (through Ovid), Embase, and CENTRAL. We won’t consider any timeframe, language, or geographical restrictions.

**Methods:** We will include only randomized controlled trials (RCTs) that have evaluated the effects of DMTs on black holes mean volume in adult patients diagnosed with any phenotype of multiple sclerosis (MS) in comparison to the placebo, routine care, or no treatment regimen. We will assess the risk of bias in the primary studies using the Revised Cochrane risk-of-bias tool for randomized trials (RoB2). Data will be synthesized based on the random-effects model and results will be plotted on a forest plot. Heterogeneity will be assessed using I2 statistics. If feasible, we will also perform subgroup analyses for each DMT.

**Funding:** This study is not funded.

**Registration:** PROSPERO submission ID: 262883.

## Introduction

### Rationale

Black holes (BH) are hypointense areas in cerebral T1 weighted magnetic resonance imaging (MRI) which represent axonal loss, neural degeneration, disease activity, or tissue destruction in patients with multiple sclerosis (MS). These lesions are called acute black holes (ABH) when an enhancement takes place simultaneously and are considered as persisting (PBH) when after the enhancement is finished (Bagnato et al., 2003).

Although neuronal destruction and axonal loss demonstrate irreversible disability in MS patients, due to the challenges of doing regular follow-up MRI to monitor BH volume progression in patients, there has not been a comprehensive evaluation on the influence of approved disease-modifying therapies (DMT) on the size of these lesions (Sahraian et al., 2010). Nevertheless, some studies have evaluated the effect of DMTs like interferon ß-1b and Glatiramer acetate on BH size in follow-up MRI images with interesting findings (Filippi et al., 2011).

This systematic review aims to evaluate the efficacy of DMTs on BH size in MS patients to present an explicit summary of the findings up to this date.

### Objectives

To evaluate the effects of FDA approved disease-modifying therapies (DMTs) on T1 hypointense lesions (Black Holes) volume in patients with multiple sclerosis (MS)

## Methods

Design and methods used for this protocol comply with Centre for Reviews and Dissemination (CRD’s) Guidance For Undertaking Reviews in Healthcare (Centre for Reviews and Dissemination, 2009) and is reported in line with Preferred Reporting Items for Systematic Reviews and Meta-Analyses Protocols (PRISMA-P) (Moher et al., 2015).

### Eligibility criteria

**(P) Population:** adult patients diagnosed with any phenotype of multiple sclerosis (MS) based on the McDonald criteria (McDonald et al., 2001) or Definite MS based on the Poser criteria (Poser et al., 1983).

**(I) Index:** all disease-modifying therapies (DMTs) that are approved by FDA, at any dose, frequency, or administration route. Concomitant interventions are allowed if they were used equally in all intervention groups in the trial. These include: Beta-1a interferon, Beta-1a peginterferon, Glatiramer acetate, Fumaric acid dimethyl ester, Teriflunomide, Fingolimod, Siponimod, Ozanimod, Natalizumab, Alemtuzumab, Ocrelizumab, Rituximab, Daclizumab, Mitoxantrone, Cladribine, and Diroximel fumarate.

**(C) Comparator:** placebo, routine care, or no treatment regimen

**(O) Outcome:** BH lesion mean volume on cerebral MRI

**(T) Timing:** to reduce the impact of confounding factors, we won’t consider the outcome in case it was measured within a considerable time gap after the initiation of the intervention. We define a considerable time gap as 6 months at maximum.

**(S) Setting:** inpatient or outpatient

We will include only randomized controlled trials (RCTs), including parallel-group randomized trials, cluster-randomized trials, and cross-over randomized trials.

### Information sources

#### Online databases

The search will employ sensitive topic-based strategies designed for each database with no time frame limitations. There will be no language or geographical restrictions either. We will perform our search on the 20^th^ of March, 2021.

Databases:

⍰ MEDLINE through Ovid Ill
⍰ Embase
⍰ Cochrane Central Register of Controlled Trials

#### Citation searching

We also examined the forward and backward citations of the included studies using Scopus.

#### Search strategy

Our search strategies for all the databases included in our study, namely MEDLINE (through Ovid), Embase, and CENTRAL are presented in Appendix A. Our search will include highly sensitive search filters for clinical trials from the InterTASC Information Specialists’ Sub-Group (ISSG) (Glanville et al., 2021) and Cochrane Collaboration (Higgins et al., 2019). To be more specific, we use a filter developed by Glanville 2019 for finding clinical trials in Embase and a filter developed by Lefebvre 2008 for the same purpose in Ovid MEDLINE. We will not apply filters to our search in CENTRAL.

### Study records

#### Data management

Records will be managed through EndNote version X9; specific software for managing bibliographies.

#### Selection process

Two reviewers (AV and MM) will independently screen the title and abstract of identified studies for inclusion. We will link publications from the same study to avoid including data from the same study more than once. If any study cannot be clearly excluded based on its title and abstract, its full text will be reviewed. A study will be included when both reviewers independently assess it as satisfying the inclusion criteria from the full text. A third reviewer (MF) will act as arbitrator in the event of disagreement following discussion. We will prepare a flow diagram of the number of studies identified and excluded at each stage following the PRISMA flow diagram of study selection (Page et al., 2020).

#### Data collection process

Using a standardized form, two reviewers (AV and MM) will extract the data independently. We will resolve any disagreements by discussion or, if required, by consultation with a third review author (MF). We will attempt to extract data presented only in graphs and figures whenever possible but will include such data only if two reviewers independently obtain the same result. If studies are multi-center, then where possible we will extract data relevant to each.

We have decided to use endpoint data and only use change-from-baseline data if the former is not available. We will also combine endpoint and change-from-baseline data in the analysis, as we want to use mean differences (MDs) (Higgins et al., 2019).

In the case of missing data, if possible, we will try to contact the original investigators to request missing information. In case of any missing data, we will try to contact the authors to receive information. In case that was unsuccessful, we will use data where attrition for a continuous outcome is between 0% and 25%, and data only from people who complete the study to that point are reported. If the standard deviation (SD) of the endpoint data was missing, we will try to impute it by standard methods using the available data (Higgins et al., 2019). That means, in case standard error (SE) was available, we will impute the SD using the formula SD = SE * √. In case confidence intervals (CI) were available, we will use the formula SD = (√ * (upper limit - lower limit) / 2Z score (if the sample size was less than 60, we will use t-score instead of Z-score). In cases where P-values for differences in means were available, first, we will calculate the corresponding t-score with the degree of freedom (df) df = Number of participants in the intervention group (n_e_) + Number of participants in the comparator group (n_c_) – 2. Then we will convert the t-score to SE using the formula SE = |MD / t|. Finally, we will convert the SE to SD using the formula 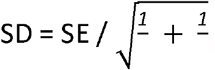

#### Data items

Data extracted will include the following summary data: sample characteristics, sample size, study methods, inclusion and exclusion criteria, MRI settings used, founding sources, declarations of interests, and results.

#### Outcomes and prioritization

Our main outcome of interest is the mean difference (MD) of T1 hypointense lesion volume after receiving DMTs between the intervention and comparator groups.

#### Risk of bias in individual studies

Two review authors (AR and MS) will assess the risk of bias for each included study. In case of disagreement between the two, a third author (MF) will act as arbitrator. We will also calculate Cohen’s kappa to assess the agreement between the two main bias assessors. We will use the Revised Cochrane risk-of-bias tool for randomized trials (RoB2) for this purpose. The tool, alongside the conditions to meet the answer “yes” for each signaling question in our review is presented in Appendix B. This tool consists of five domains: bias arising from the randomization process, bias due to deviations from the intended interventions, bias due to missing outcome data, bias in the measurement of the outcome, and bias in the selection of the reported results. This tool is specifically designed for parallel-groups randomized controlled studies. To assess the risk of bias in the clustered randomized controlled trials and cross-over randomized controlled trials, we will use two variants of this tool specifically designed for such studies. These three tools are identical in all domains, except that the formers also assess the randomization methods specific for cluster or cross-over. Using these three tools alongside each other does not introduce any methodological deficiency. The answer for each signaling question can be: yes, probably yes, probably no, no, and no information provided. We will judge the overall risk of bias for each domain as either high, some concerns, or low according to the manual of the tools.

#### Data synthesis

We will use the R version 4 (Team, R.C, 2013) “meta” package (Schwarzer, 2007) and “rob.summary” (Harrer et al., 2019) package as the software for our data synthesis. We expect our outcome of interest to be reported in means (μ) after receiving the intervention for both the intervention and comparator group. Because these outcomes are expected to be reported in the same unit (millimeters), we will use mean differences (MDs) for the statistical analysis. We will calculate the variance and standard error of those MDs. Because of the nature of our intervention of interest, we expect some variability in the studies. Thus, we will perform a meta-analysis on those values based on the random-effects model. We seek our effect of interest as the effect of assignment to the intervention (Intention-to-treat effect). If the authors applied such a strategy, we will use their results. If the original authors presented only the results of the per-protocol, we will assume that those participants lost to follow-up would have had the same percentage of events as those who remained in the study.

#### Cluster trials

If authors fail to account for intra-class correlation in clustered studies, it will lead to a unit-of-analysis error whereby P values are spuriously low, CIs unduly narrow, and statistical significance overestimated (Divine et al., 1992). Where clustering has been incorporated into the analysis of primary studies, we will present these data as if from a non-cluster randomized study but will adjust for the clustering effect. If cluster studies have been appropriately analyzed and intra-class correlation coefficients and relevant data documented in the report taken into account, synthesis with other studies will be possible using the generic inverse variance technique. We will try to contact the first authors of studies to obtain intra-class correlation coefficients for their clustered data and to adjust for this (Gulliford et al., 1999). In case we couldn’t account for the intra-class correlation coefficients, we will use the standard methods reported by Higgins et al. (Higgins et al., 2019) and will mark these studies in the analysis with an asterisk (*).

#### Cross-over trials

The major concern of cross-over trials is the carry-over effect. This means that the treatment’s effect in the first phase is carried over to the second phase (Elbourne et al., 2002). As this is very likely for DMTs, we will only use data from the first phase of cross-over studies.

### Other analyses

#### Assessment of heterogeneity

We will inspect our data visually to investigate the possibility of statistical heterogeneity. We will also perform I^2^ statistics alongside the Chi^2^ P-value (Deeks, 2001). I^2^ statistic quantifies inconsistency across studies to assess the impact of heterogeneity on the meta-analysis (Higgins & Thompson, 2002). Chi^2^ statistics will be considered substantial if there was a low P-value (less than 0.10). We planned to interpret the I^2^ statistic as follows:

⍰ 0% to 40%: might not be important;
⍰ 30% to 60%: may represent moderate heterogeneity;
⍰ 50% to 90%: may represent substantial heterogeneity;
⍰ 75% to 100%: represents considerable heterogeneity.

If we find moderate (or more) heterogeneity, we will attempt to determine possible reasons for it by subgroup analyses.

#### Subgroup analyses

If at least 5 studies are available for each DMT, we will perform a subgroup analysis for it. Reasons for heterogeneity in the outcome will also be explored by the restricted-maximum-likelihood-random-effect meta-regressions.

#### Sensitivity analyses

We will undertake a sensitivity analysis testing how prone the outcome is to change when data only comes from the studies that presented the data as intention-to-treat analysis, compared with when we also have studies with the effect of adhering to the intervention (Per-protocol effect) analyses in our final analysis.

In case we had cluster trials that we couldn’t account for the intra-class correlation coefficients in, we will perform a sensitivity analysis on trials that have no the same issue to test how prone the outcome is to change when data is not affected by unit-of-analysis issues.

If we had to impute the missing information for primary studies, we will examine the validity of the imputations in a sensitivity analysis that excludes imputed values.

We will also analyze the effects of excluding trials that are judged to be at high risk of bias across one or more of the “Risk of bias” domains.

#### Meta-bias

To evaluate the risk of reporting bias across studies, a contour-enhanced funnel plot alongside a test for funnel plot asymmetry will be conducted. Contour lines will correspond to perceived ‘milestones’ of statistical significance (P = 0.01, 0.05, and 0.1). These contours may help differentiate asymmetry due to non-reporting biases from that due to other factors. If studies appear to be missing in areas where results would be statistically non-significant and unfavorable to the experimental intervention then this adds credence to the possibility that the asymmetry is due to non-reporting biases. If the supposed missing studies are in areas where results would be statistically significant and favorable to the experimental intervention, this would suggest the cause of the asymmetry is more likely to be due to factors other than non-reporting biases. The test for funnel plot asymmetry examines whether the relationship between estimated effect size and study size is greater than chance (Higgins et al., 2019).

#### Confidence in cumulative evidence

The strength of the overall body of evidence will be assessed using the Grading of Recommendations, Assessment, Development and Evaluation (GRADE) framework (GRADE Working Group, 2004), which takes into account seven criteria: Risk of bias, Consistency of effect, Imprecision, Indirectness, and Publication bias. Two review authors (MS and AR) will rate the certainty of the evidence for the outcome as ‘high’, ‘moderate’, ‘low’, or ‘very low’. We will resolve any discrepancies by consensus, or, if needed, by arbitration by a third review author (MF).

## Data Availability

Important protocol amendments post registration will be recorded and included in dissemination.

## Administrative information

### Registration

Submitted in the International Prospective Register of Systematic Reviews (PROSPERO) under the submission number: 262883.

### Contributions

MF is the leading author for protocol development, analyses, and dissemination. MF is also the first reviewer. AV is the corresponding author. All authors will contribute to data interpretation and article drafts.

### Amendments

Important protocol amendments post registration will be recorded and included in dissemination.

### Support

This study is not funded.

## Appendices

## Appendix A Search Strategies

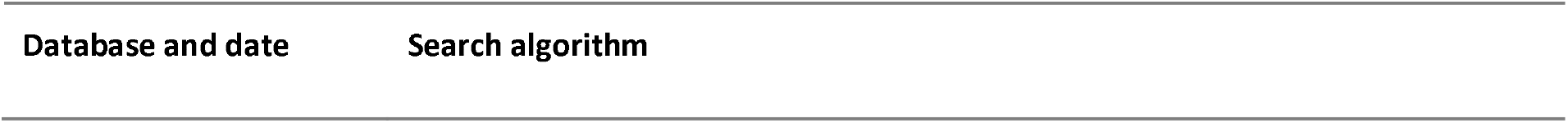
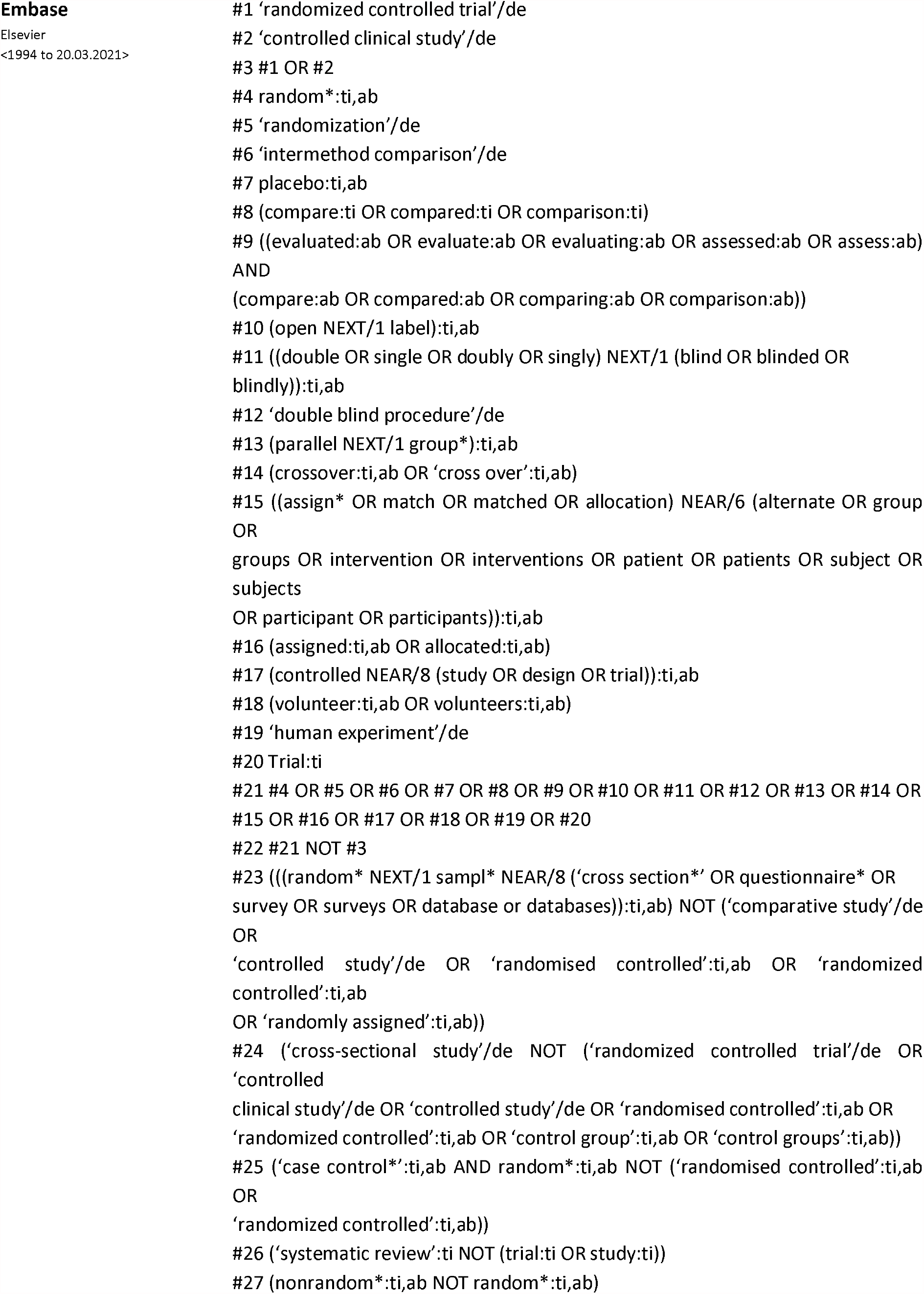
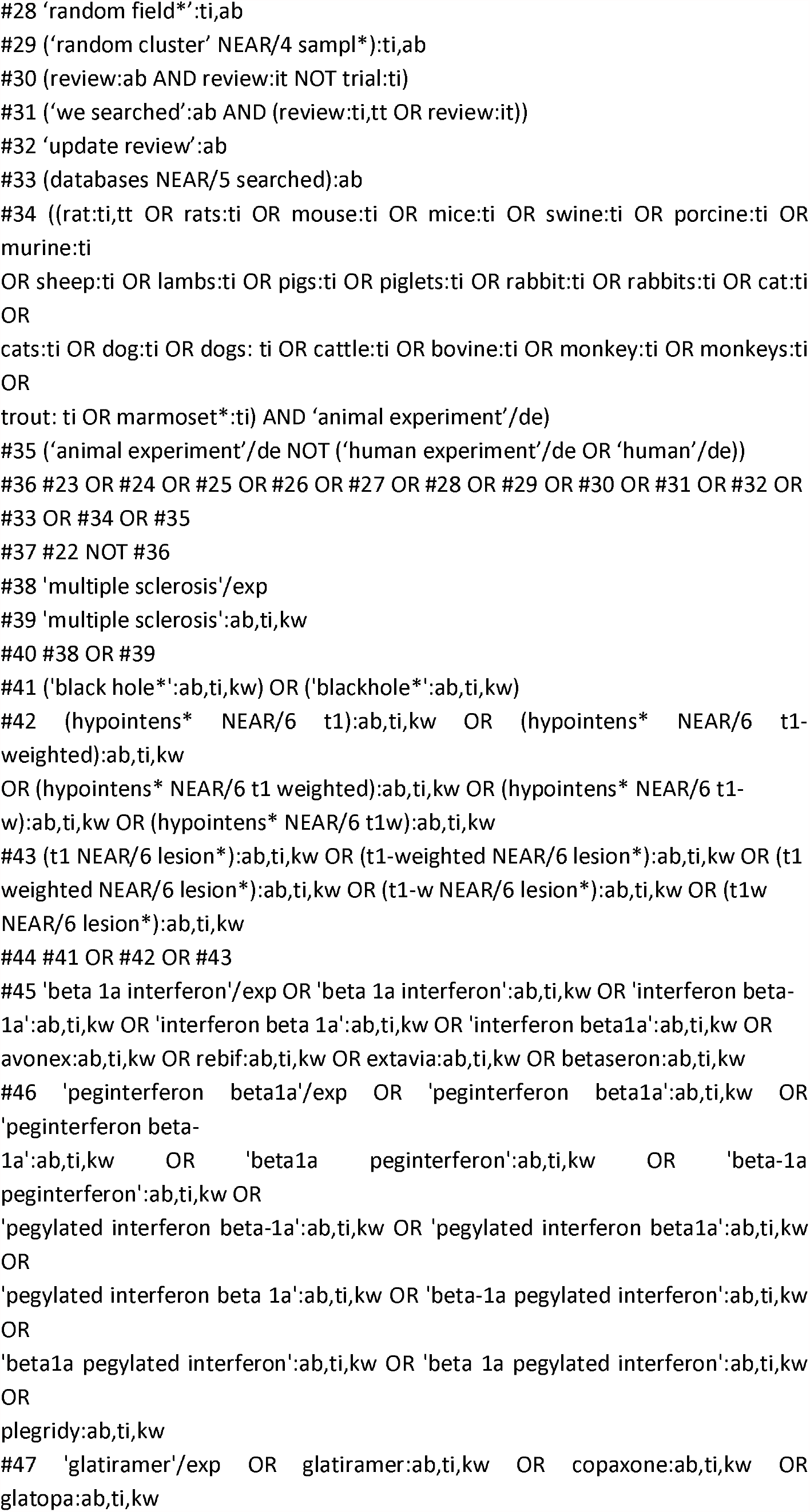
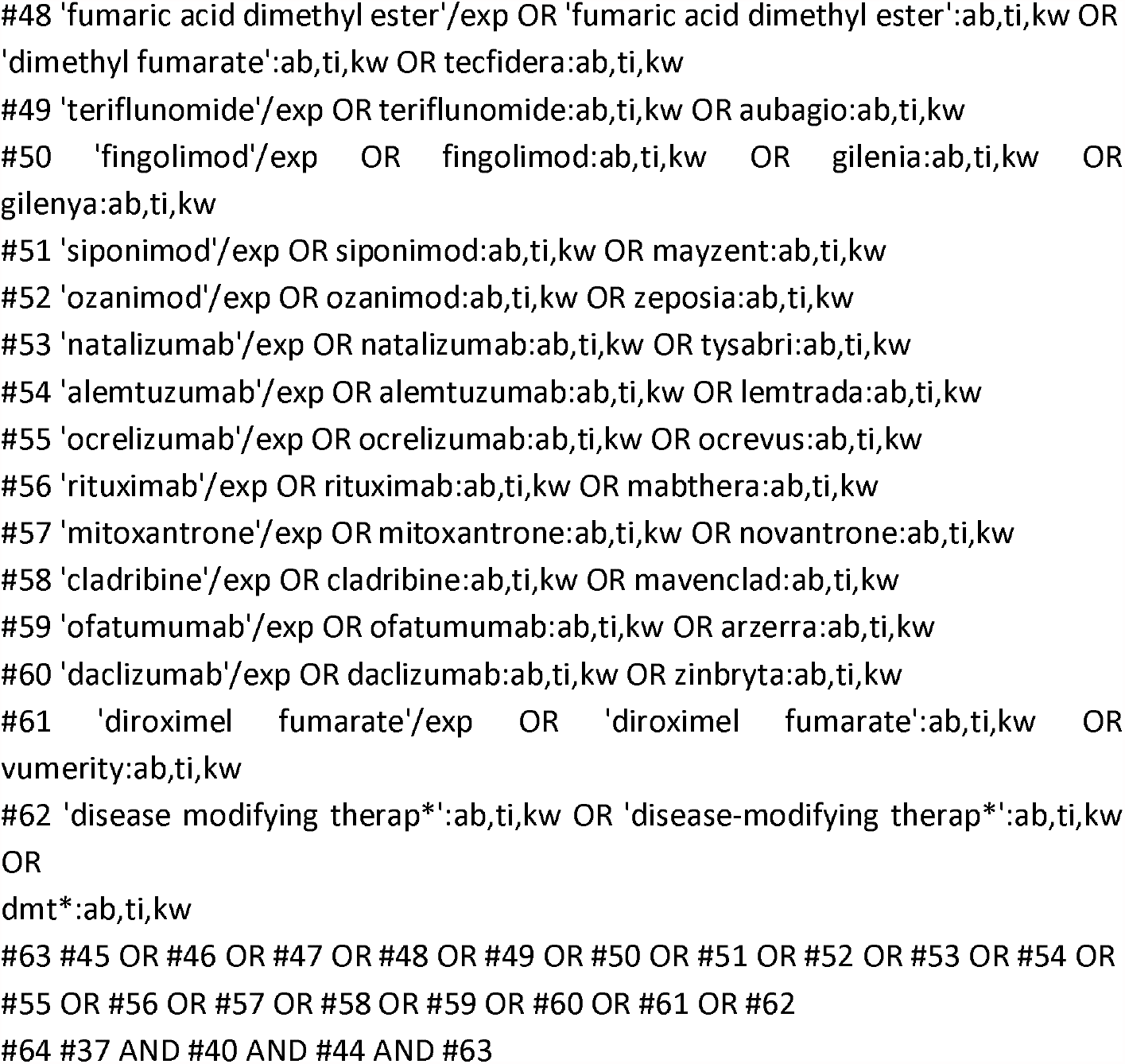
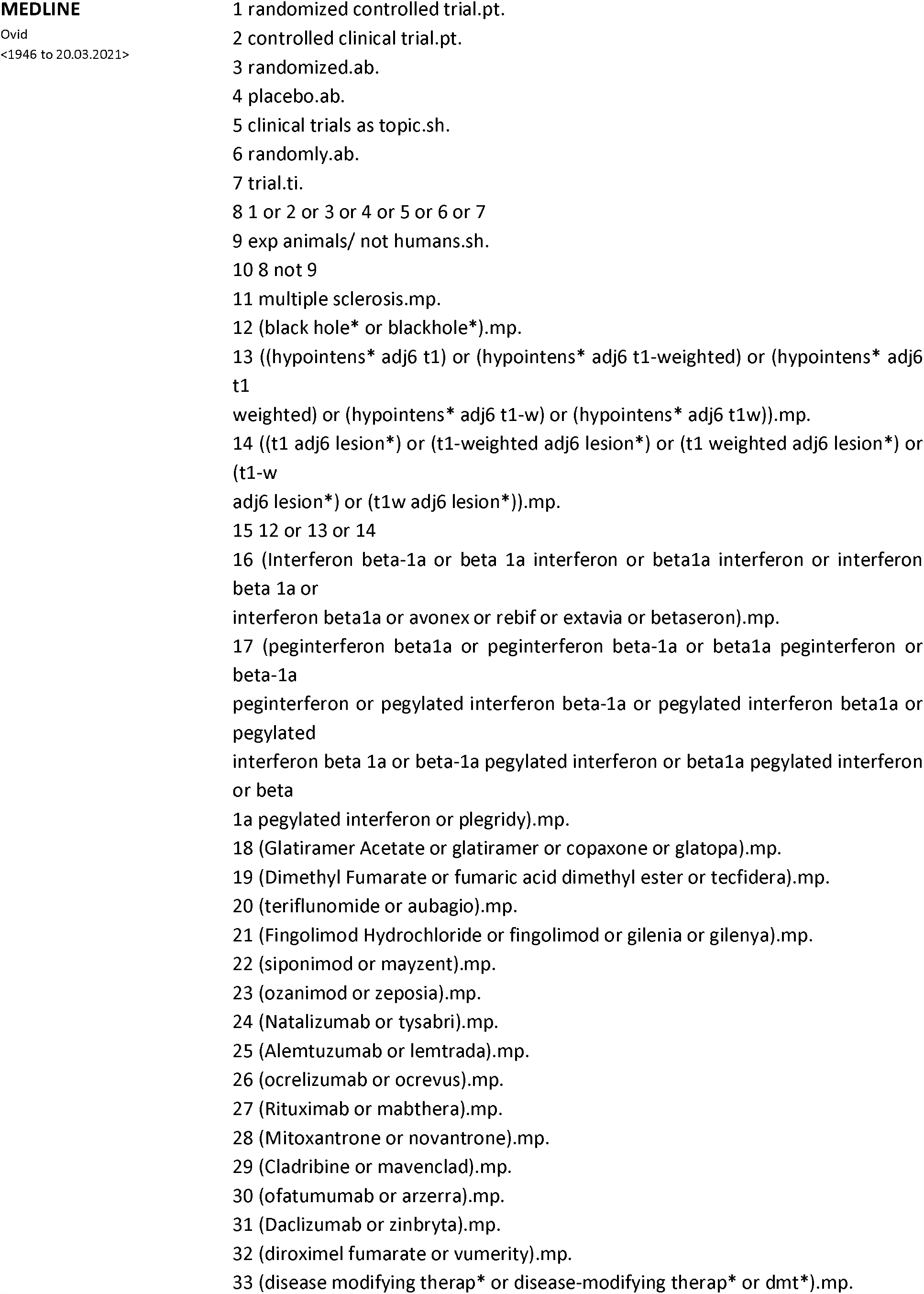
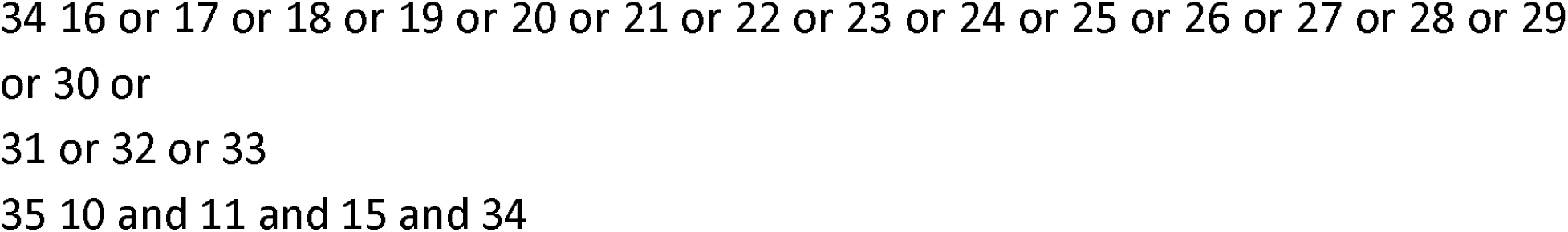
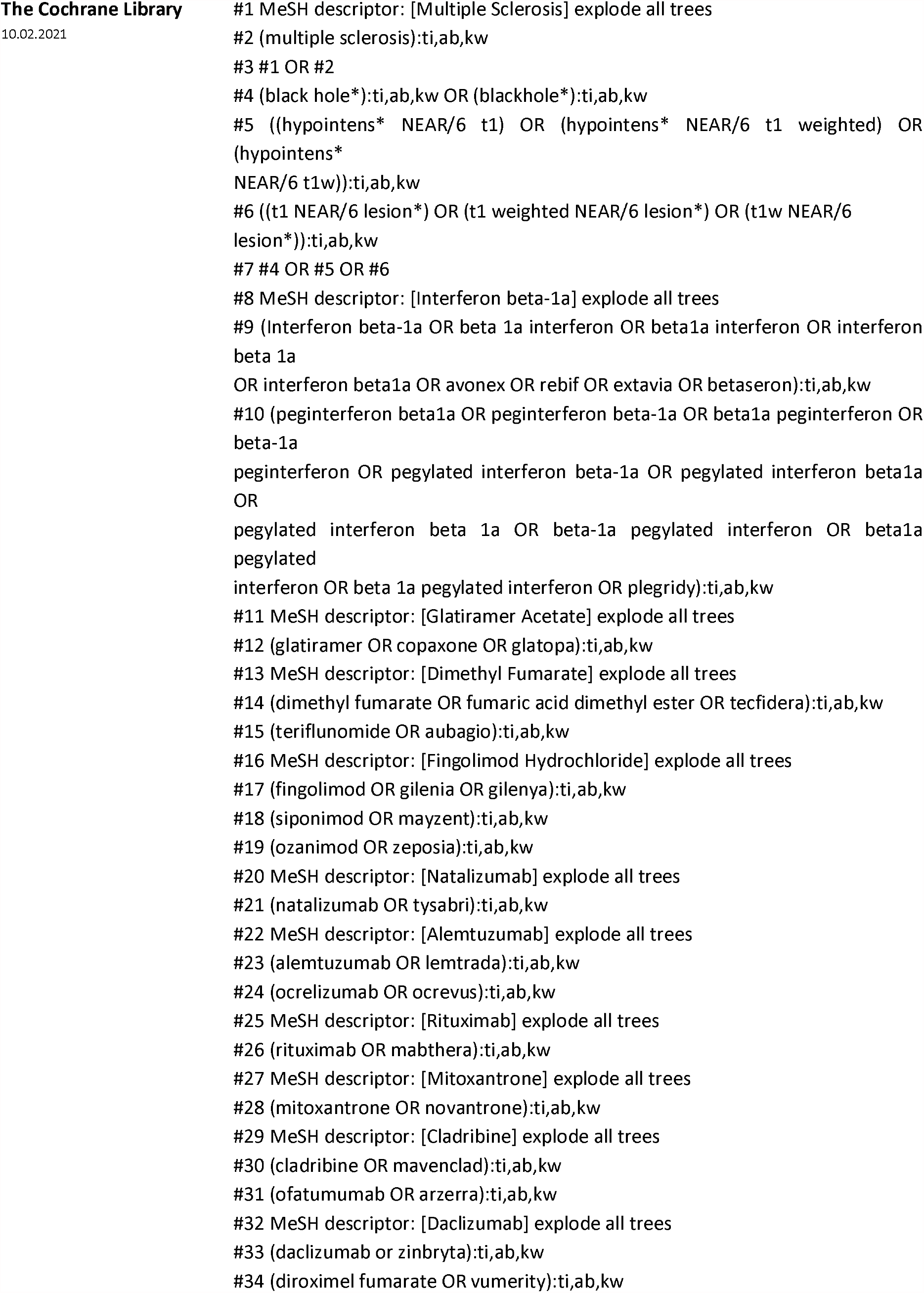
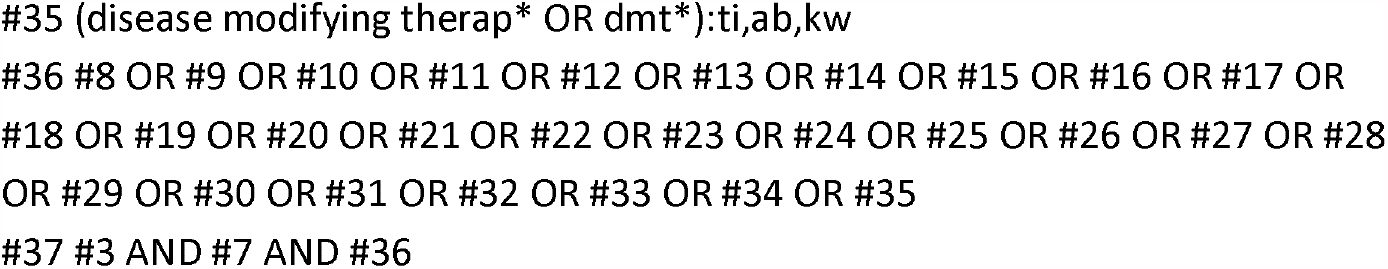

## Appendix B, The RoB2 tool and its variants

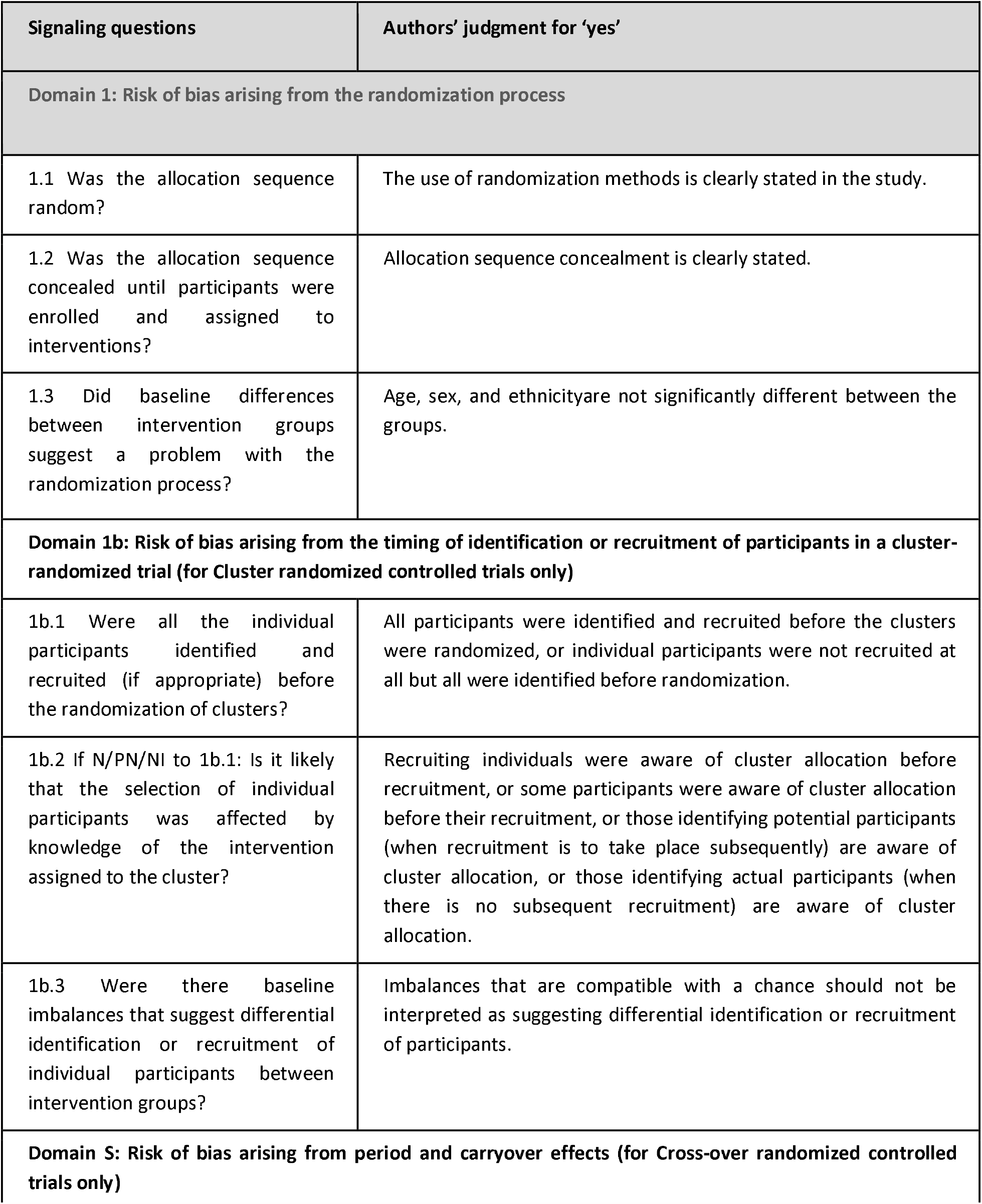
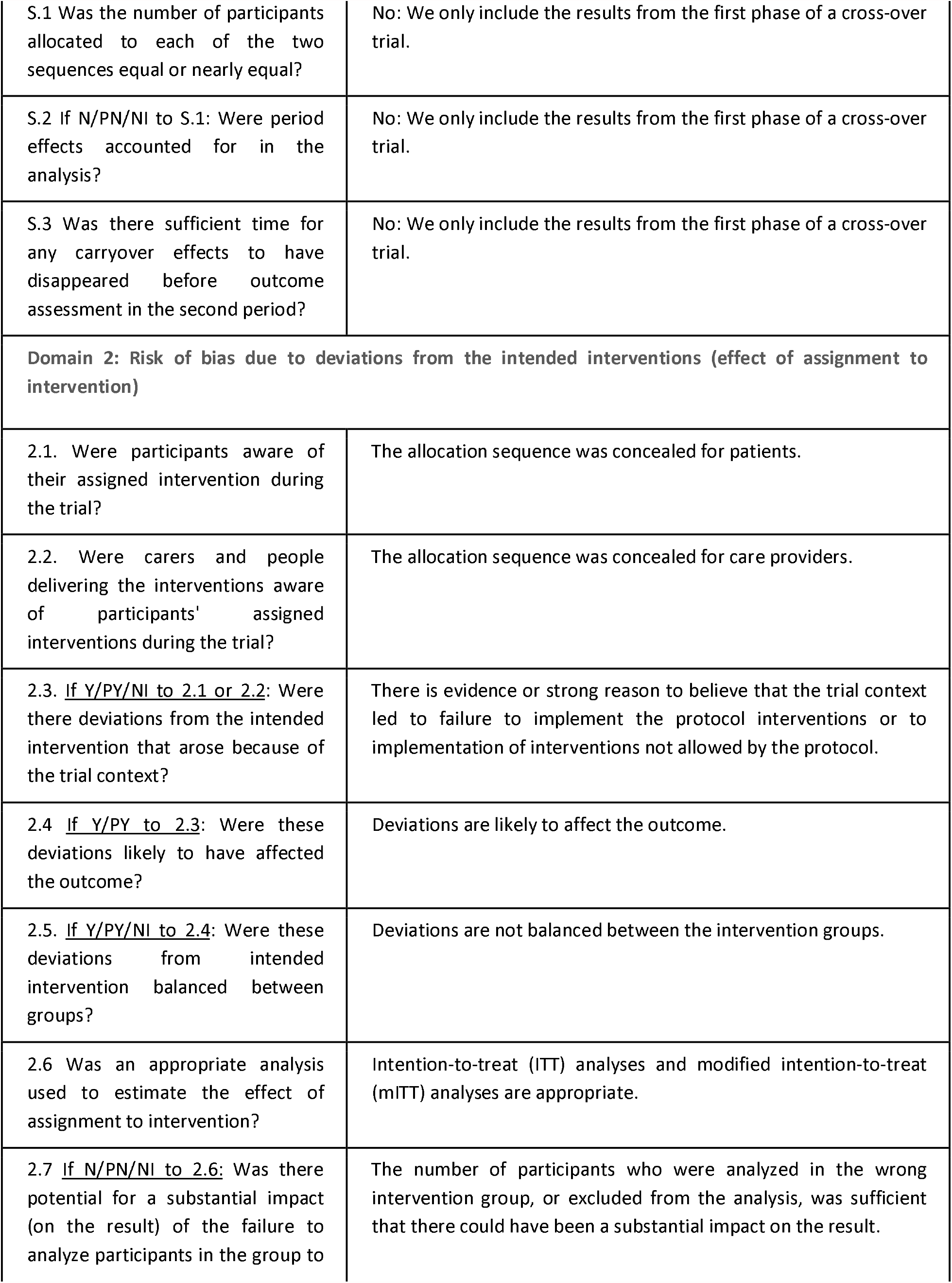
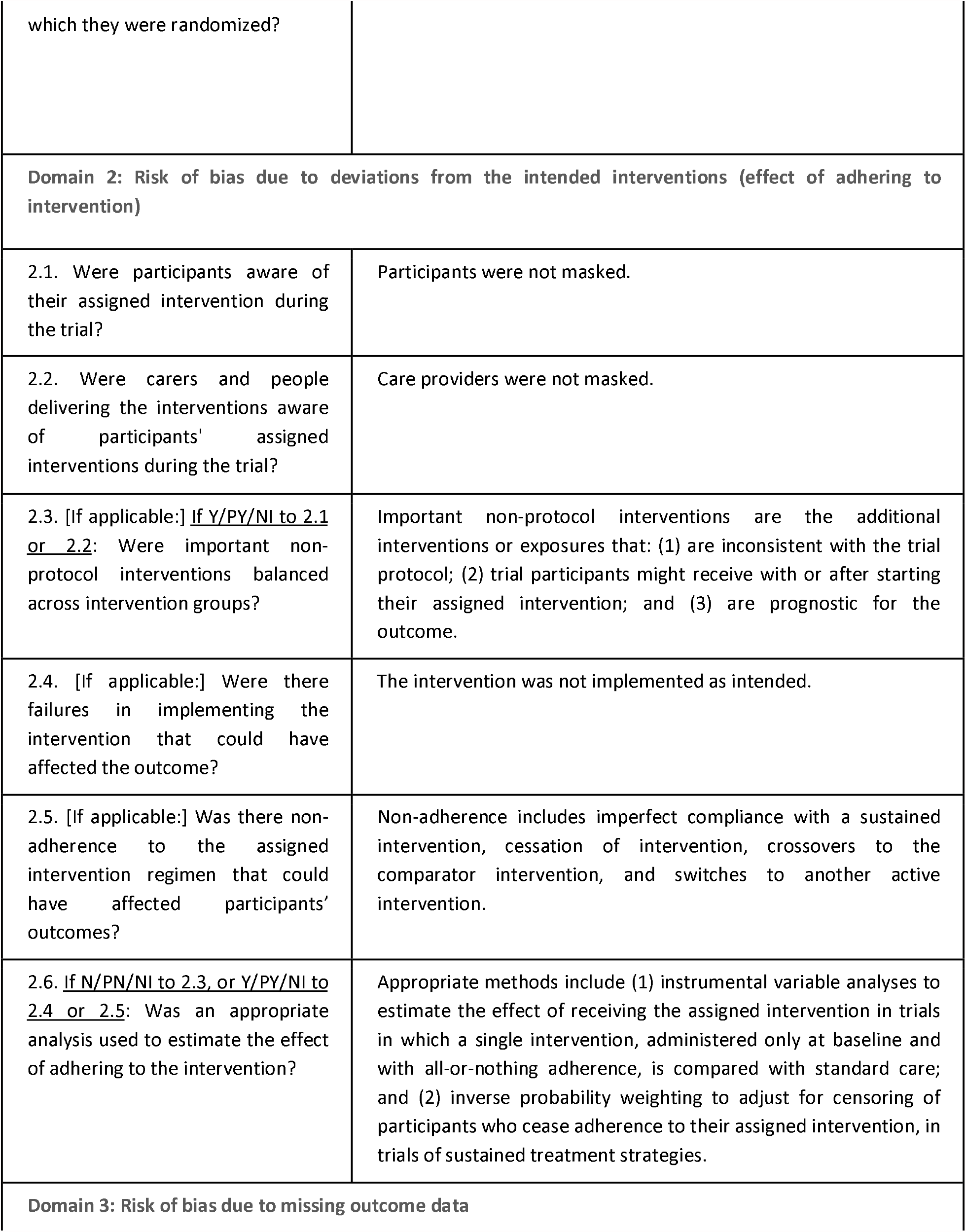
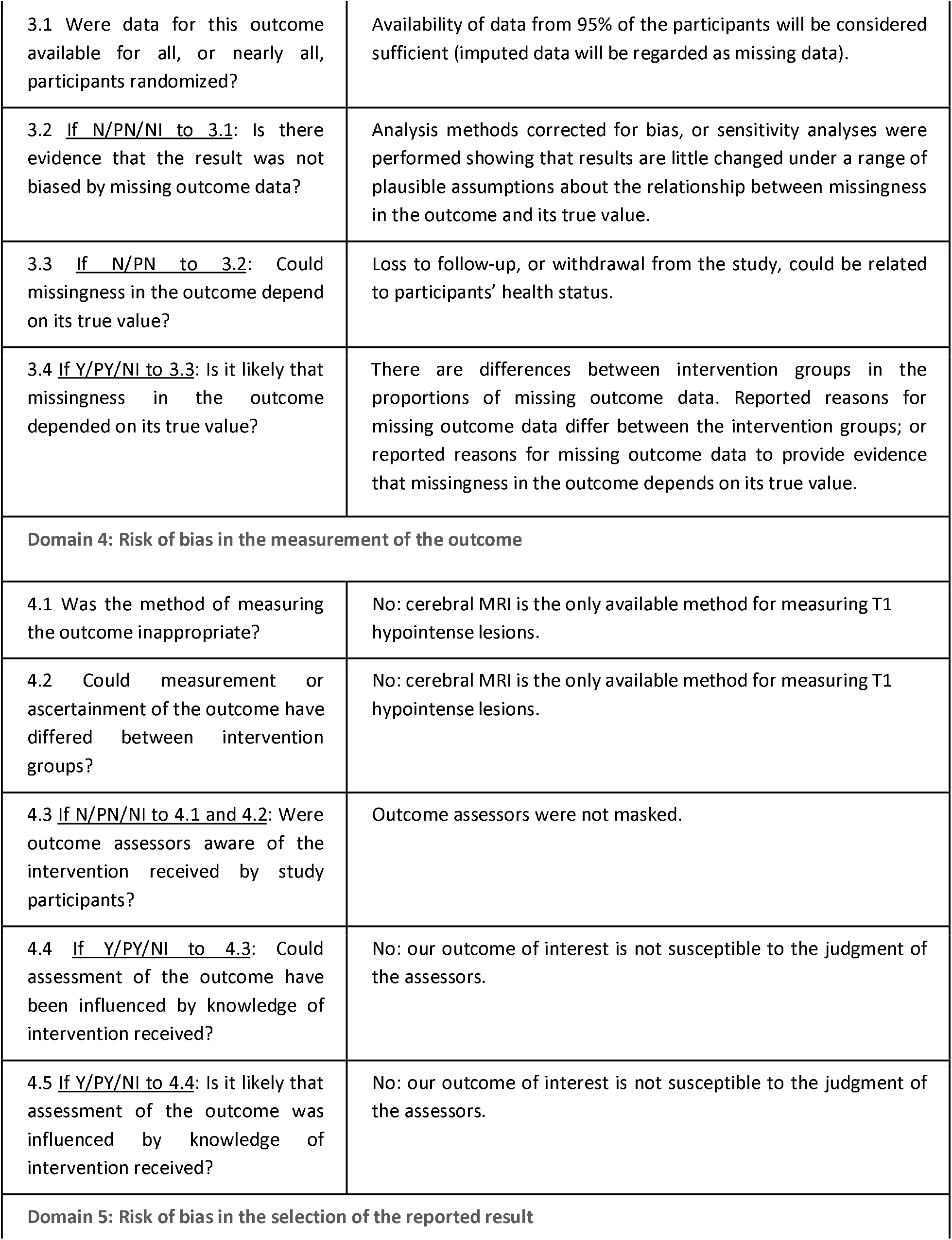
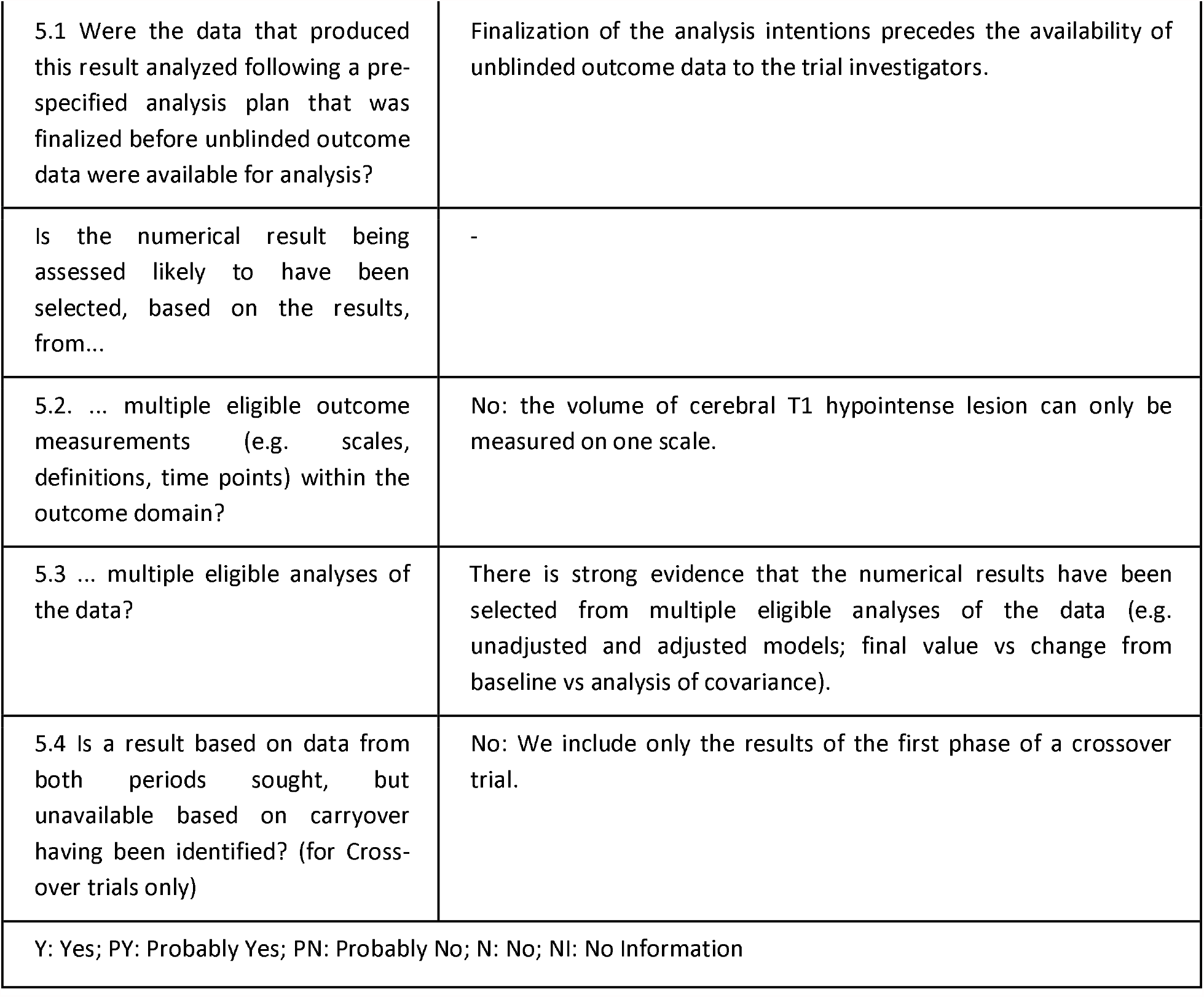

